# Early Phylogenetic Estimate Of The Effective Reproduction Number Of 2019-nCoV

**DOI:** 10.1101/2020.02.19.20024851

**Authors:** Alessia Lai, Annalisa Bergna, Carla Acciarri, Massimo Galli, Gianguglielmo Zehender

**Affiliations:** Department of Biomedical and Clinical Sciences “L.Sacco”, University of Milan, Milano, Italy; CRC-Coordinated Research Center “EpiSoMI”, University of Milan, Milano, Italy; Romeo ed Enrica Invernizzi Pediatric Research Center

## Abstract

To reconstruct the evolutionary dynamics of the 2019 novel coronavirus, 52 2019-nCOV genomes available on 04 February 2020 at GISAID were analysed.

The two models used to estimate the reproduction number (coalescent-based exponential growth and a birth-death skyline method) indicated an estimated mean evolutionary rate of 7.8 × 10^−4^ subs/site/year (range 1.1×10^−4^–15×10^−4^).

The estimated R value was 2.6 (range 2.1-5.1), and increased from 0.8 to 2.4 in December 2019. The estimated mean doubling time of the epidemic was between 3.6 and 4.1 days.

This study proves the usefulness of phylogeny in supporting the surveillance of emerging new infections even as the epidemic is growing.

## INTRODUCTION

On 30 January 2020, the World Health Organisation (WHO) declared that the outbreak of an infection due to a novel coronavirus (2019-nCoV) was a “Public Health Emergency of International Concern” (https://www.who.int/news-room/detail/30-01-2020-statement-on-the-second-meeting-of-the-international-health-regulations-(2005)-emergency-committee-regarding-the-outbreak-of-novel-coronavirus-(2019-nCoV)). Emerging as a human pathogen in the Chinese city of Wuhan, 2019-nCoV (https://www.who.int/docs/default-source/coronaviruse/situation-reports/20200121-sitrep-1-2019-ncov.pdf?sfvrsn=20a99c10_4) has caused a widespread outbreak of febrile respiratory illness and, as of 13 February 2020, there were 60,349 confirmed cases (including 527 outside mainland China) and a total of 1,360 fatalities (https://gisanddata.maps.arcgis.com/apps/opsdashboard/index.html#/bda7594740fd40299423467b48e9ecf6).

Belonging to the β*-coronavirus* genus of the *Coronaviridae* family, 2019-nCoV is closely related to SARS-CoV as there is >70% nucleotide similarity in their approximately 30 kb long genomes.^1^ A recent study has supported the view that, like other *β-coronaviruses* causing human infections such as SARS-CoV and MERS-CoV, 2019-nCoV originated from bats, and reported 96% genomic identity with a previously detected SARS-like bat coronavirus.^2,3^ However, it remains unclear whether the spillover also involved a different intermediary animal host.

In the case of such an epidemic, it is important to make an as reliable as possible estimate of the basic reproductive number (R^0^, the number of cases generated from a single infected person) and the dynamics of transmission. The aim of this study was to investigate the temporal origin, rate of viral evolution and population dynamics of 2019-nCoV using 52 full genomes of viral strains sampled in different countries on known sampling dates available at the moment when study was performed.

## MATERIALS AND METHODS

### Sequence data set

The analysis was based on 52 2019-nCOV sequences publicly available at GISAID (Global Initiative on Sharing All Influenza Data) on 4 February 2020 (https://www.gisaid.org/). The accession IDs, sampling dates and locations are summarized in Table S1.

The sequences were aligned using the ClustalW Multiple Alignment program included in the accessory application of Bioedit software, manually controlled, and cropped to a final length of 29,774 bp using BioEdit v. 7.2.6.1 (http://www.mbio.ncsu.edu/bioedit/bioedit.html).

### Phylodynamic analysis

The simplest evolutionary model best fitting the sequence data was selected using software JmodelTest v.2.1.7 software,^4^ and proved to be the Hasegawa-Kishino-Yano (HKY) model.

The virus’ phylogeny, evolutionary rates, times of the most recent common ancestor (tMRCA) and demographic growth were co-estimated in a Bayesian framework using a Markov Chain Monte Carlo (MCMC) method implemented in v.1.84 of the BEAST package.^5^

Different coalescent priors and molecular clock models (constant population size, exponential growth, and a Bayesian skyline plot, BSP) were tested using strict and relaxed molecular clock models. Given the large credibility interval and high level of uncertainty due to very close sampling dates, all the estimates were made using days as the unit of time and a normal prior with substitution rates obtained from our preliminary estimates (mean rate 2.2 × 10^−6^ subs/site/day, with a standard deviation of 1.1 × 10^−6^).

The MCMC analysis was run until convergence with sampling every 100,000 generations. Convergence was assessed by estimating the effective sampling size (ESS) after 10% burn-in using Tracer v.1.7 software (http://tree.bio.ed.ac.uk/software/tracer/), and accepting ESS values of 300 or more. The uncertainty of the estimates is indicated by 95% highest marginal likelihoods estimated^6^ by path sampling/stepping stone methods.^7^

The final trees were summarised by selecting the tree with the maximum product of posterior probabilities (pp) (maximum clade credibility or MCC) after a 10% burn-in using Tree Annotator v.1.84 (included in the BEAST package), and were visualised using FigTree v.1.4.2 (http://tree.bio.ed.ac.uk/software/figtree/).

The basic reproductive number (R_0_) was calculated on the basis of the exponential growth rate (r) using the equation R_0_=rD+1, where D is the average duration of infectiousness estimated as described below.^8^ The doubling time of the epidemic was directly estimated setting the tree prior to the coalescent exponential growth analysis with doubling time parameterization.

### Birth-Death Skyline estimates of the effective reproductive number (R_e_)

The birth-death skyline model implemented in Beast 2.48 was used to infer changes in the effective reproductive number (R_e_), and other epidemiological parameters such as the death/recovery rate (δ), the transmission rate (λ), the origin of the epidemic, and the sampling proportion (ρ).^9^ Given that the samples were collected during a short period of time, a “birth-death contemporary” model was used.

The analyses were based on the previously selected HKY substitution model and the substitution rate was set to the value of 8.0 × 10^−4^ subs/site/year, which corresponds to the mean substitution rate estimated using a relaxed clock under the exponential coalescent model as transformed into units per year.

For the birth-death analysis, one and two intervals and a log-normal prior for R_e_, with a mean (M) of 0.0 and a variance (S) of 1.0 were chosen, which allows the R_e_ values to change between <1 (0.193) to >5. A normal prior with M=48.7 and S=15 (corresponding to a 95% interval from 24.0 to 73.4) was used for the rate of becoming uninfectious. These values are expressed as units per year and reflect the inverse of the time of infectiousness (5.3-19 days, mean 7.5) according to the serial interval estimated by Qun Li *et al*.^10^ Sampling probability (ρ) was estimated assuming a prior Beta (alpha=1.0 and beta=999), corresponding to a minority of the sampled cases (between 10^−5^ to 10^−3^). The origin of the epidemic was estimated using a normal prior with M=0.1 and S=0.05 in units per year.

The MCMC analyses were run for 30 million generations and sampled every 3,000 steps.

Convergence was assessed on the basis of ESS values (ESS >200). Uncertainty in the estimates was indicated by 95% highest posterior density (95%HPD) intervals.

The mean growth rate was calculated on the basis of the birth and recovery rates (r=λ-δ), and the doubling time was estimated by the equation: doubling time=ln(2)/r.^11^

## RESULTS

The sequence analyses under a relaxed (uncorrelated log-normal) or strict molecular clock showed that the former performed better as assessed by using BF with path sampling (PS) and stepping stone sampling (SS) (strict *vs*. relaxed molecular clock BF(PS)=-8.66 and BF(SS)=-10.7 for relaxed clock). Comparison of the different demographic models showed that the BSP model best fitted the data (BSP *vs*. exponential growth BF(PS)= 7.3 and BF(SS)= 8.78 for BSP; BSP *vs*. constant population size BF(PS)= 7.3and BF(SS)= 8.78 for BSP). The estimated mean evolutionary rate was 2.15 × 10^−6^ subs/site/day (95% HPD: 3.22 × 10^−7^–4.18 × 10^−6^), corresponding to 7.8 × 10^−4^ subs/site/year (95% HPD: 1.1 × 10^−4^–15 × 10^−4^).

The estimated mean tMRCA corresponding to the root of the tree dated 73 days before the end of January 2020 (95%HPD: 32.5–142.3), corresponding to 18 November 2019 (95%HPD: 10 September 2019-28 December 2019).

The Bayesian tree showed three main significant clades. The largest clade (pp=0.84) encompassed 10 sequences and consisted of two significant sub-clades (pp=0.9 and pp=1). Overall, this cluster included fewer recent isolates than the other two clusters, and dated back to 47.5 days ago (95% HPD: 25.5-76.6), corresponding to 13 December 2019. The second (pp=0.99) and third significant clusters (pp=0.95) dated back to 29.2 (95% HPD: 0.7-47.45) and 21.9 (95% HPD 3.6-54.7) days ago, corresponding to 01-08 January 2020.

The Bayesian skyline plot showed a rapid increase in the number of infections in a period between about 45 and 30 days before the end of January 2020 (Fig.1, part A).

The IDs and available data of the sequences involved in the clades are shown in Table S1.

The estimated growth rate under the exponential growth model was 0.218 days^-1^, corresponding to an R_0_ estimation of 2.6 (credibility interval: 2.1-5.1). The direct estimation of the doubling time of the epidemic gave a mean 3.6 days (varying from 1.0 to 7.7). Figure 1 (part B) shows the Bayesian birth-death skyline plot of the R_e_ estimates with 95%HPD, and indicates that R_e_ increased from <1 (mean 0.8, 95%HPD: 0.3-1.3) to a mean value of 2.4 (95%HPD: 1.5-3.5) in December 2019, and has since remained at this value.

**Fig. 1.**
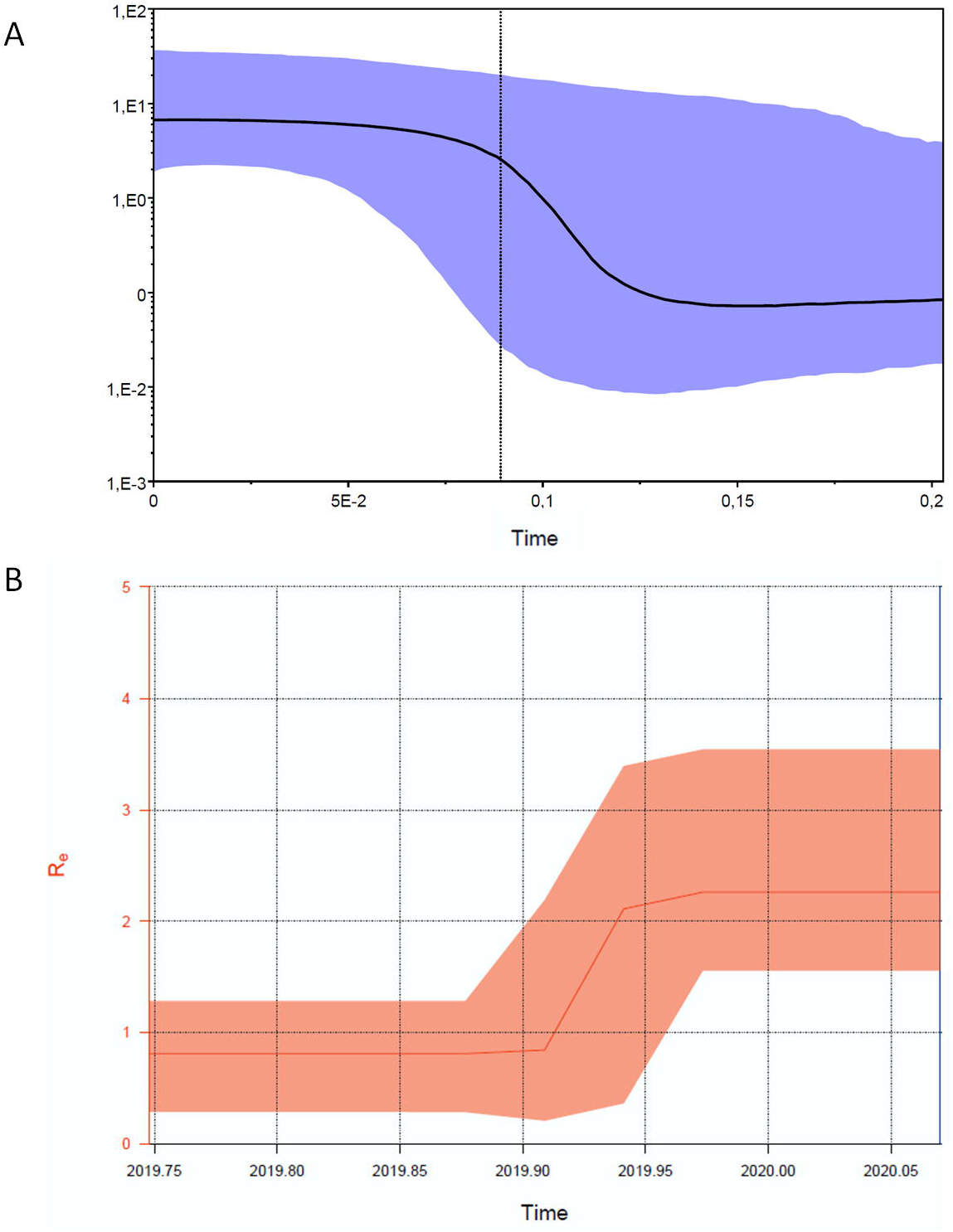
Part A: Bayesian Skyline plot of the 2019-nCoV outbreak. The Y axis indicates Ne and X axis shows the time in year units (0=January 30; 0.05=18.2 days; 0.1=36.5 days; 0.15=54.7 days and 0.2=73 days before). The thick solid line represents the median value of the estimates, and the grey area the 95% HPD. Part B: Birth-death skyline plot of the 2019-nCoV outbreak allowing two Re intervals. The curve and the orange area show the mean Re values and their 95% confidence intervals. The Y and X axes respectively represent R values and time in years.

Table 1 shows the parameters estimated using the birth-death skyline plot. The epidemic originated an estimated mean 3.7 months (credibility interval 3-4) before the present (BP), corresponding to October-November 2019, before the root tree (3.6 months BP). The estimated recovery rate (the time to becoming non-infectious) was 7.3 days (CI 4.7-16.5 days), whereas the transmission rate (λ) increased from 40.5 to 112.4 in units per year in December 2019. On the basis of these values, the growth rate in the second period is r=0.17 (0.16-0.19), corresponding to a mean doubling time of 4.1 days (range 3.9-4.3).

**Table 1.**
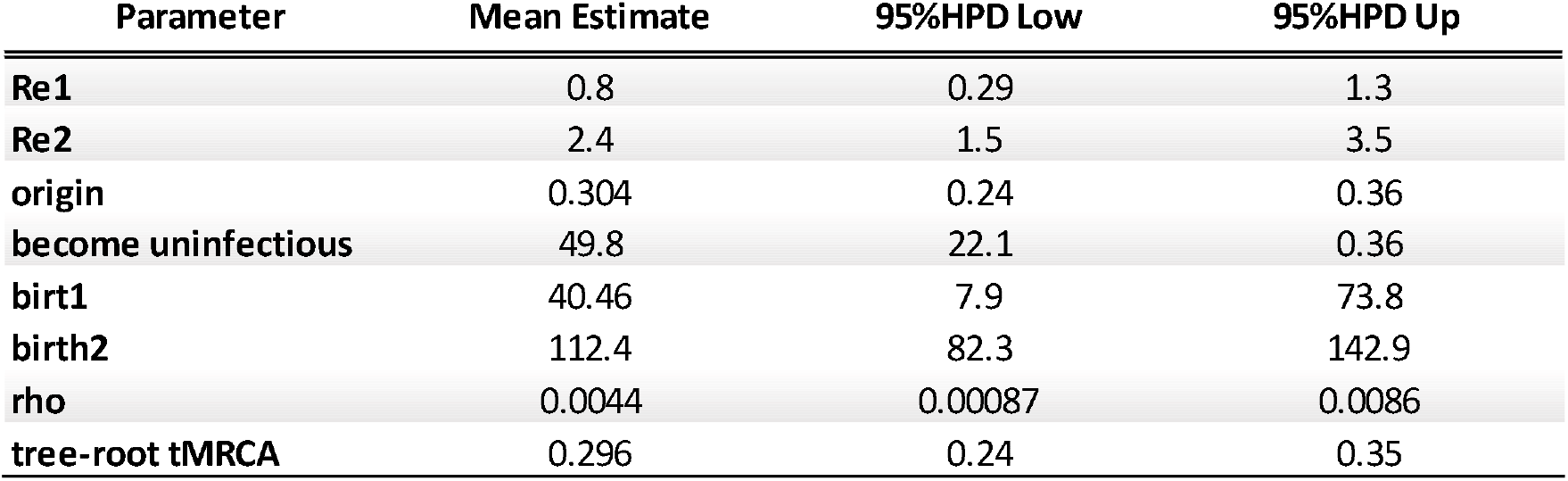
Epidemiological parameters estimated by Birth-death skyline analysis.

## DISCUSSION

The 2019-nCoV epidemic is unique in the history of human infectious diseases not only because it is caused by a novel virus, but also because of the immediate availability of epidemiological and genomic data (the first entire genome was published on 24 December 2019). The prompt availability of research data on internet platforms such as the GISAID has allowed us and other research groups to make a phylogenetic reconstruction of the origin of 2019-nCoV and to share these findings with other scientists.

The temporal reconstruction of the 2019-nCoV phylogeny obtained in the present study is in line with previous estimates and suggests that the epidemic originated between October and November 2019, several weeks before the first cases were described. This was confirmed by means of coalescent analysis and the birth-death method of estimating the origin of the epidemic. The estimated evolutionary rate is also in line with that of SARS and MERS viruses,^12,13^ and the recent estimates concerning 2019-nCoV (http://virological.org/t/phylodynamic-analysis-67-genomes-08-feb-2020/356).

One of the most important epidemiological parameters when monitoring an epidemic is R_0_ (i.e. the number of secondary cases induced by a single infected individual in a totally susceptible population) because it is fundamental to assess the potential spread of a micro-organism. Its value changes during an epidemic being called the effective reproduction number (Re). R_0_ is usually estimated on the basis of the growth rate of the number of cases. The available epidemiological estimates of 2019-nCoV R_0_ range from 2.2 to 2.9, although they changed from 1.4 to >7 during the first phases of the epidemic.^10,14^ Recently developed evolutionary models have made it possible to estimate epidemiological parameters on the basis of phylogenesis,^9,15^ and a coalescent and a birth-death methods were used to estimate R_0_ and the changes in the R_e_ of the 2019-nCoV epidemic during a short period of time. This has allowed us to make a preliminary estimate that mean R_0_ from the beginning of the epidemic to the first days of February 2020 was 2.2 (range 3.6-5.8), and the birth-death skyline analysis showed an increase in R_e_ from <1 to 2.4 (CI 1.5-3.5) during December 2019. This agrees with the BSP analysis showing an increase in the number of infections in the same period of time.

On the same basis, the estimated epidemic doubling time was 3.6 days with a credibility interval between 1 to 7 days. We also tried to calculate it on the basis of the transmission (λ) and recovery rate (δ) estimated using the birth-death model, which lead to an estimated mean doubling time of 4.1 days, with the most probable values falling between 3.9 and 4.3 days. Previous studies have suggested that the doubling time during the early phases of the epidemic was about 7.4 days.^10^ The difference in the estimate here obtained, may be due to the increased epidemic growth rate observed during the last days of January, or the initial delay in recognising and reporting new cases.

This preliminary study has some limitations. The R values and doubling times were estimated phylogentically using all of the whole genomes available in a public database at the time the study was carried out (https://www.gisaid.org/). Given the small number of sequences and the relatively short sampling period, the credibility intervals are wide and limit the precision of the estimates. Moreover, the analysis included isolates collected outside mainland China as it is assumed that they all belong to the same epidemic originating in Wuhan.

Serial intervals were used to estimate the duration of infectiousness, although we do not yet have any information concerning the possible existence and duration of a latent (pre-infectious) period that would contribute to the serial interval.

More detailed and accurate analyses can be made when a larger number of genomes and more precise data on the infectious period become available. However, although the R_0_ calculated on the basis of the direct observation of the number of infected individuals may be affected by omissions or delayed notifications of cases,^16^ a phylogenetic estimate of the same parameter may be more reliable.

This became particularly evident recently (on February 12, 2020) when the change in diagnosis classification led to a sudden increase in the reported cases by Hubei, China (https://myemail.constantcontact.com/COVID-19-Updates---Feb-12.html?soid=1107826135286&aid=Kdg8a0rBTAk).

In conclusion, these results allowed us to make a phylogenetic estimate of the R_0_ of 2019-CoV infection that is similar to that obtained using conventional epidemiological methods^17^ (https://www.who.int/news-room/detail/23-01-2020-statement-on-the-meeting-of-the-international-health-regulations-(2005)-emergency-committee-regarding-the-outbreak-of-novel-coronavirus-(2019-ncov), and a possibly shorter estimated doubling time of the number of subjects involved at least during the early phases of the epidemic. They also support the usefulness of phylodynamic as an important complement to classic approaches to the surveillance and monitoring of an emerging infection, even during the course of an epidemic.

## Data Availability

All sequences used are publicly available at GISAID(Global Initiative on Sharing All Influenza Data)website

https://www.gisaid.org/

## ACKNOWLEDGEMENTS

In memory of Li Wenliang, Carlo Urbani and of all the doctors and health workers who endangered their lives in the fight against epidemics.

We acknowledge the authors, originating and submitting laboratories of the sequences from GISAID.

